# Impact of COVID-19 vaccine doses and viral waves on inflammatory and immunological responses to COVID-19 infections in India

**DOI:** 10.1101/2024.12.19.24319150

**Authors:** Kamal Kant Sharma, Uttara Partap, Yogesh Marathe, Sanaa Shaikh, Pradeep D’Costa, Gaurav Gupta, Molin Wang, Wafaie W Fawzi, Kevin C Kain, Nerges Mistry, Yatin Dholakia

## Abstract

**Background:** Investigation of the effect of SARS-CoV-2 variants and COVID-19 vaccination on inflammatory and immune response to SARS-CoV-2 infection is limited in South Asia.

**Objectives:** We aimed to examine the impact of COVID-19 vaccination and waves of COVID- 19 on inflammatory and immunological biomarkers among COVID-19 patients in India.

**Methods:** This cross-sectional analysis used baseline data from a randomized controlled trial of vitamin D and zinc during COVID-19 infection in India (N=181). Blood samples and data regarding vaccination doses were collected. The second (Delta) or third (Omicron) wave was determined by date of enrolment. Mixed effects linear regression with robust standard errors was used to examine associations between COVID-19 vaccination dose or wave at enrolment and C-Reactive Protein (CRP), ferritin, lactate dehydrogenase (LDH), D-dimer, interleukin-6 (IL-6), angiopoietin-2 (Ang-2), soluble triggering receptor expressed on myeloid cells-1 (sTREM-1), immunoglobulin G (IgG) and immunoglobulin M (IgM).

**Results:** Compared to no vaccination, full vaccination was associated with lower LDH (*P<*0.001), D-dimer (*P=*0.521) and Ang-2 (*P=*0.046), and higher IgG levels (*P<*0.001). Partial vaccination was associated with lower IL-6 (*P=*0.040) and higher IgG (*P<*0.001). Enrolment during the third wave was associated with lower IL-6 (*P<*0.001), CRP (*P=*0.056), IgM (*P=*0.013), and IgG (*P<*0.001), but higher D-dimer levels (*P<*0.001).

**Conclusions:** COVID-19 vaccination status and SARS-CoV-2 variant influence the inflammatory and immunologic response during SARS-CoV-2 infection, contributing to the severity of clinical presentation.

## Introduction

COVID-19 infection is caused by the severe acute respiratory syndrome coronavirus 2 (SARS- CoV-2), a major global health concern of the 21st century, and has affected more than 609 million people and caused over 6.5 million deaths worldwide.^[1]^ The ability of SARS-CoV-2 viruses to undergo frequent and random mutations may result in the development of highly contagious and lethal variants, and this has extended the COVID-19 pandemic through multiple waves of infection.^[1]^

The progression of COVID-19 infection is associated with altered clinical manifestations and changes in the level of related inflammatory and immunological biomarkers. Innate immune responses, coagulation, and the initiation of pro-inflammatory responses are common pathways to counteract infectious agents, which may result in increased circulating levels of acute phase proteins such as C-reactive protein (CRP), ferritin, and D-Dimer. In particularly acute cases, the proliferation of the virus in the host cell and its rapid spread in other cells results in a cytokine storm, which is considered a critical condition.^[2]^ The profile of human immune response, transmission ability, laboratory characteristics, and antigenicity of the virus depends at least partially on the COVID-19 virus variant.^[3,4]^ While differences in symptoms have been noted,^[5]^ there are limited data available on inflammatory and immunological responses to different SARS-CoV-2 variants, particularly from South Asian populations.

Protective immunity through vaccines was paramount to reducing the severity of COVID-19 disease and breaking the chain of transmission. India also scaled up its domestic vaccine research and development program for the launch of the vaccination drive in the country by January 2021 and has achieved over 2 billion vaccinations so far.^[6]^ Around 921 million people had their first dose of the COVID-19 vaccine in India, 863 million were fully vaccinated (2 doses), and around 139 million received their precautionary dose by September 2022.^[7]^ It has been observed that vaccination and its doses directly impact the inflammatory and antibody response to SARS-CoV-2 infection.^[8,9]^ However, evidence regarding the effect of COVID-19 vaccination on the inflammatory and immunological response among newly-infected patients remains limited, particularly from lower- and middle-income populations such as those in South Asia.^[10]^

Few data have been reported on a comprehensive range of inflammatory and immunological markers by COVID-19 vaccination doses and second versus third waves of the COVID-19 pandemic (which roughly correspond to two distinct SARS-CoV-2 variants – Delta and Omicron respectively) from COVID-19 patients, particularly in India. In this study, we aimed to determine the association between COVID-19 vaccination doses and waves of COVID-19, and a set of inflammatory and immunological biomarkers, among SARS-CoV-2- infected patients in Maharashtra, India.

## Materials and Methods

### Study procedures

This analysis uses baseline data from a randomized controlled trial aiming to evaluate the effects of vitamin D and zinc supplementation on COVID-19 outcomes in Maharashtra, India (ClinicalTrials.gov registration: NCT04641195).^[11]^ From April 2021 to February 2022, hospitalized or home-isolated COVID-19 patients aged ≥18 years with a positive RT-PCR or rapid antigen test and with mild to moderate symptoms were enrolled from two study sites: King Edward Memorial (KEM) Hospital and Research Centre, Pune, and Saifee Hospital, Mumbai.

Details regarding study procedures have been outlined previously,^[11]^ baseline procedures are briefly described as follows. At enrolment, background information, medical history, and clinical symptoms were obtained and clinical assessment was conducted along with the collection of blood samples for routine laboratory investigations and study of specific inflammatory and immunological biomarkers. In addition, participants were asked about details of their COVID-19 vaccination status at baseline and during successive follow-ups.

Routine blood investigation included testing of CRP, lactate dehydrogenase (LDH), serum ferritin, and D-dimer. This was performed at the respective hospital sites, which were accredited by the National Accreditation Board for Testing and Calibration Laboratories (NABL). All COVID-19 treatment, such as medications, was delivered by clinicians independently of the study, guided by the national treatment recommendations for COVID-19 at the time.^[12]^

Testing of other plasma biomarkers including angiopoietin-2 (Ang2), interleukin-6 (IL-6), soluble triggering receptor expressed on myeloid cells-1 (sTREM-1), immunoglobulin G (IgG), and immunoglobulin M (IgM) were conducted at the coordinating institute, The Foundation for Medical Research (FMR). Blood samples collected were processed at hospital sites for plasma, and the centrifuged matrix was transported to FMR adhering to the transportation regulations provided by the Government of India (GOI) for COVID-19 samples.^[13]^ Aliquots of plasma were prepared for further testing and stored at −20°C. The concentration of biomarkers was quantified using sandwich enzyme-linked immunosorbent assay (ELISA), was performed according to the manufacturer’s instructions; Thermofisher Invitrogen, USA (IL-6, Ang-2, sTREM-1) and RayBiotech, USA (IgG and IgM) and readings were observed at 450 nm. Due to issues with the sample kit, sTREM-1 assays were performed only for a subset of 48 participants. To ensure accurate results, the samples were run in triplicates.

### Variables of interest

Exposures of interest were COVID-19 vaccination status and wave during which the participant was enrolled. Participants were categorized based on their COVID-19 vaccination doses received till the day of enrolment. Participants with no COVID-19 vaccine were grouped as unvaccinated, those with one dose were partially vaccinated, and those who received second dose and/or the final third (precautionary or booster) dose were categorized as fully vaccinated. Participants were enrolled across two waves of the COVID- 19 pandemic in India. These waves were defined as wave 2 (predominantly Delta; enrolled from the start of the study in April 2021 to 15^th^ December 2021), and wave 3 (predominantly Omicron; enrolled from 21^st^ December 2021, onwards – no participants were enrolled between 15^th^-21^st^ December 2021).^[14]^

Outcomes of interest were the following inflammatory and immunological biomarkers: serum CRP, LDH, and ferritin, plasma D dimer, IL-6, Ang2, sTREM-1, IgG, and IgM. As part of the analysis, we considered other potentially important participant characteristics including sex, age, education, health behaviour (smoking and alcohol consumption), pre-existing medical conditions, COVID-19 medication, body mass index (BMI), COVID-19 vaccine type, median number of days with symptoms and median days since last vaccine dose before enrolment.

### Statistical analysis

First, baseline demographic, clinical, health behaviour, COVID-19 medication, and anthropometric characteristics were compared across the two exposures of interest: 1) COVID-19 vaccination and 2) COVID-19 waves. Differences between groups were examined by Pearson’s Chi-squared tests or Fisher’s exact tests (comparisons with cell counts <5) for categorical variables, and Wilcoxon rank sum tests or Kruskal Wallis tests (comparisons across >2 groups) for continuous variables. We also examined median biomarker levels across the two exposures of interest, testing for differences across groups using Wilcoxon rank sum tests or Kruskal Wallis tests, as described above.

Following this, effects of the exposures of interest on each outcome were modelled using mixed-effects linear regression with robust standard errors. Models were adjusted for the other exposure (either COVID-19 vaccination or wave), and for age, sex, health behaviour (smoking and alcohol consumption), pre-existing medical conditions, COVID-19 medication, BMI, COVID-19 vaccine type, and median number of days with symptoms and were additionally adjusted for clustering at the hospital site level. Although we also initially included days since last vaccination in regression models, this variable was dropped due to collinearity with the COVID-19 vaccination variable. Missing observations for covariates were minimal (maximum: 2 participants [1.1%] for any covariate) and we assumed these were missing completely at random. We used the missing indicator method to handle missing data in regression models. All analyses were performed using STATA, version 14.1 (StataCorp, Texas).

### Ethics

The procedures followed in this study were in accordance with the ethical standards of all responsible ethical committees listed below, and with the Declaration of Helsinki, and informed written consent was taken from study participants. Ethical approval was obtained from the Institutional Review Board of the Harvard T.H. Chan School of Public Health (Protocol No. IRB20-1425), the University Health Network Research Ethics Board (20-5775), the Institutional Research Ethics Committee of the Foundation for Medical Research (IREC No. FMR/IREC/C19/02/2020), the Institutional Review Board of Saifee Hospital (Project No. EC/008/2020) and the KEM Hospital Research Centre Ethics Committee (KEMHRC ID No. 2027). The trial is registered on ClincialTrials.gov: NCT04641195, and Clinical Trial Registry of India (CTRI): CTRI/2021/04/032593. Health Ministry Steering Committee (HSMC) - GOI endorsement was received before the commencement of the study.

## Results

### Overall baseline and clinical characteristics of the enrolled participants

Total 181 COVID-19 patients were included in the study. The mean (SD) age of the study participants was 47 (15.7) years, and the range was 18-87 years. One-fourth of participants (24.8%) were 60 years of age or older. The proportion of male participants was 51.3%, and 91.8% of participants had received secondary or higher-level education. Around 62% of participants were overweight or obese (30.9% in each category), only 6 individuals (3.3%) were underweight and a normal BMI was found in 34.8% participants. The median (interquartile range [IQR]) number of days with symptoms before enrolment was 3 days (2- 4). 39.7% of participants reported comorbidities such as hypertension (24.8%), diabetes (20.9%), and cardiovascular disease (7.7%), and 19.8% of participants were on any COVID-19 medications. Among the antiviral medications, Remdesivir was used the most frequently (12.2%) (Table 1).

**Table 1.**
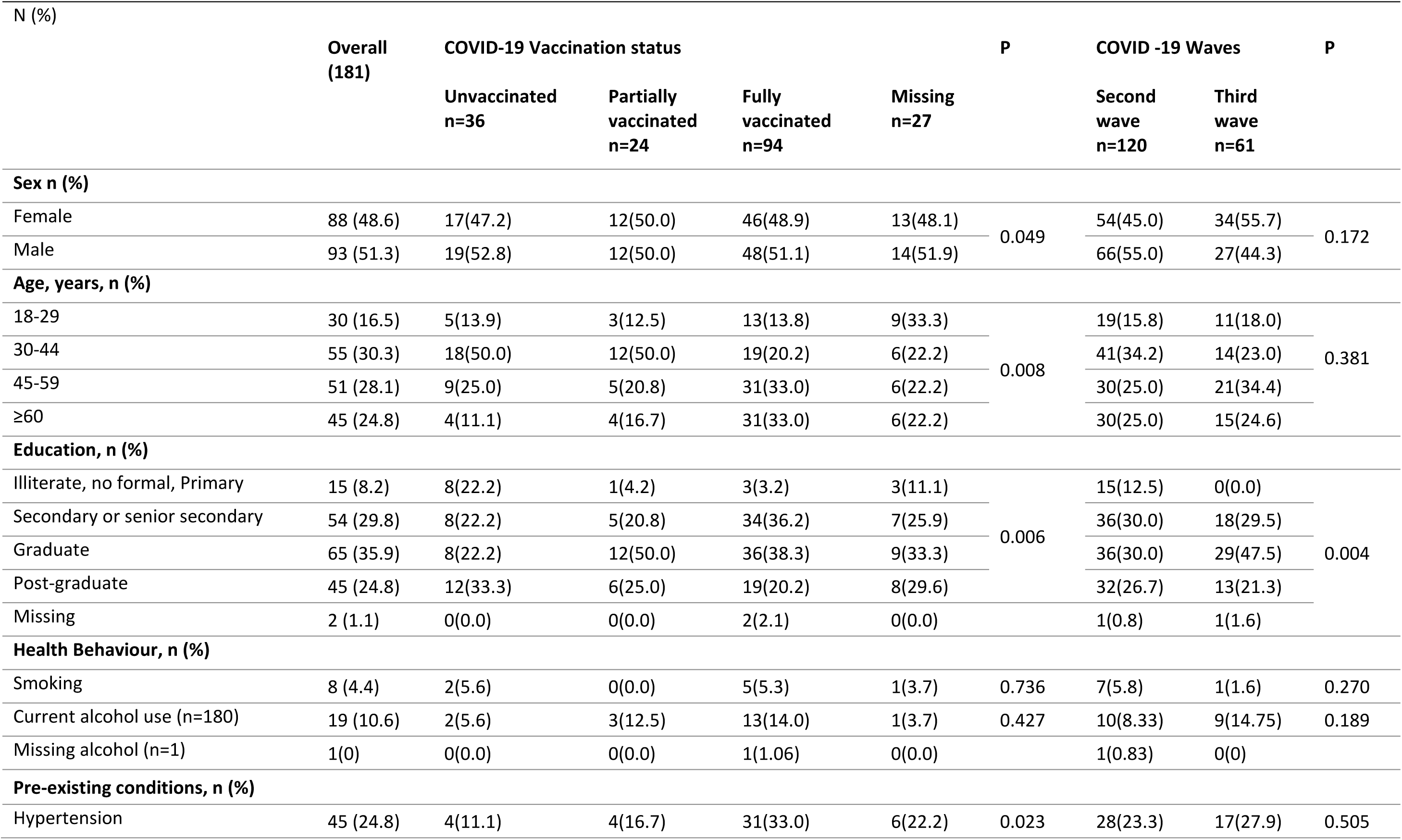
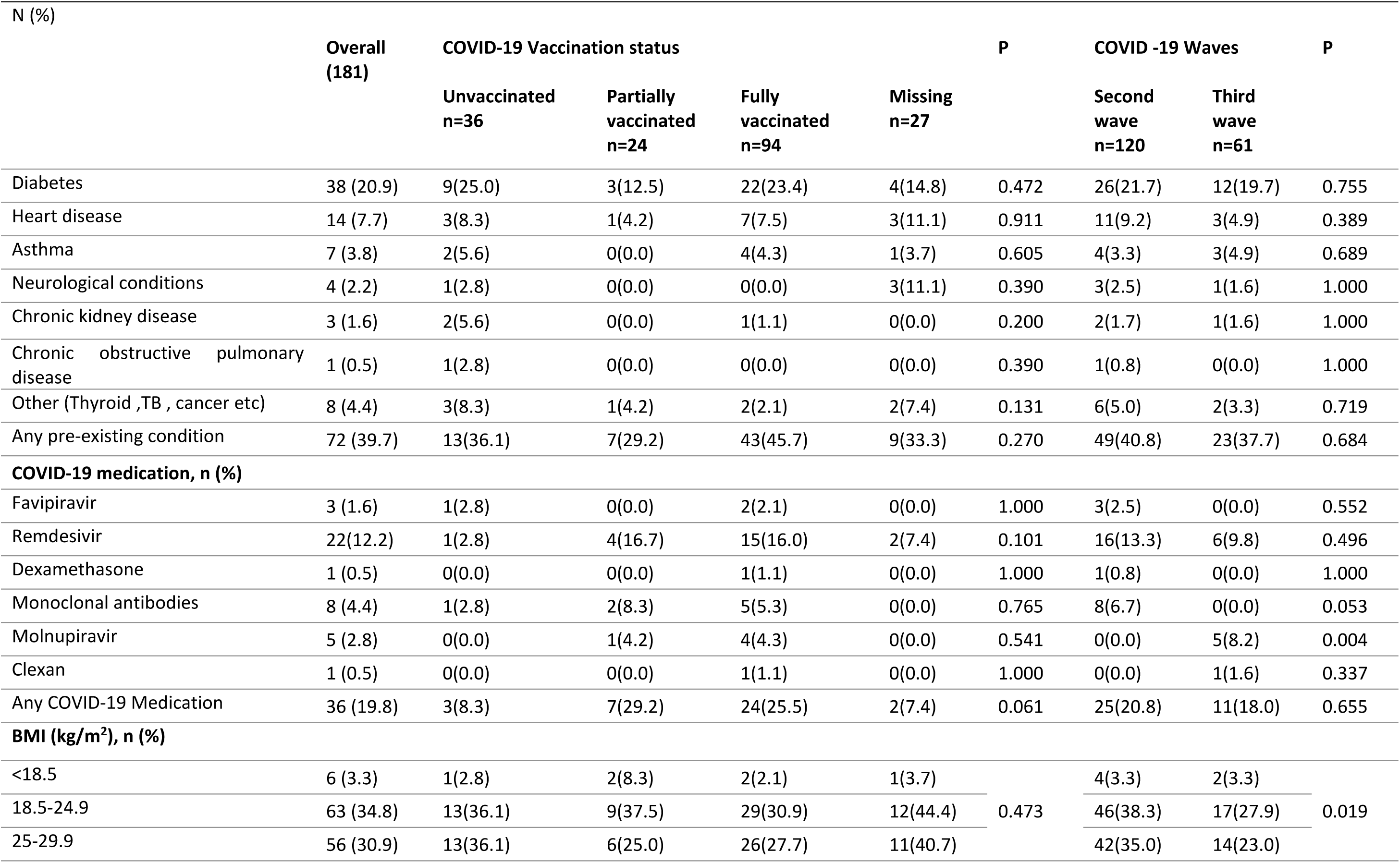
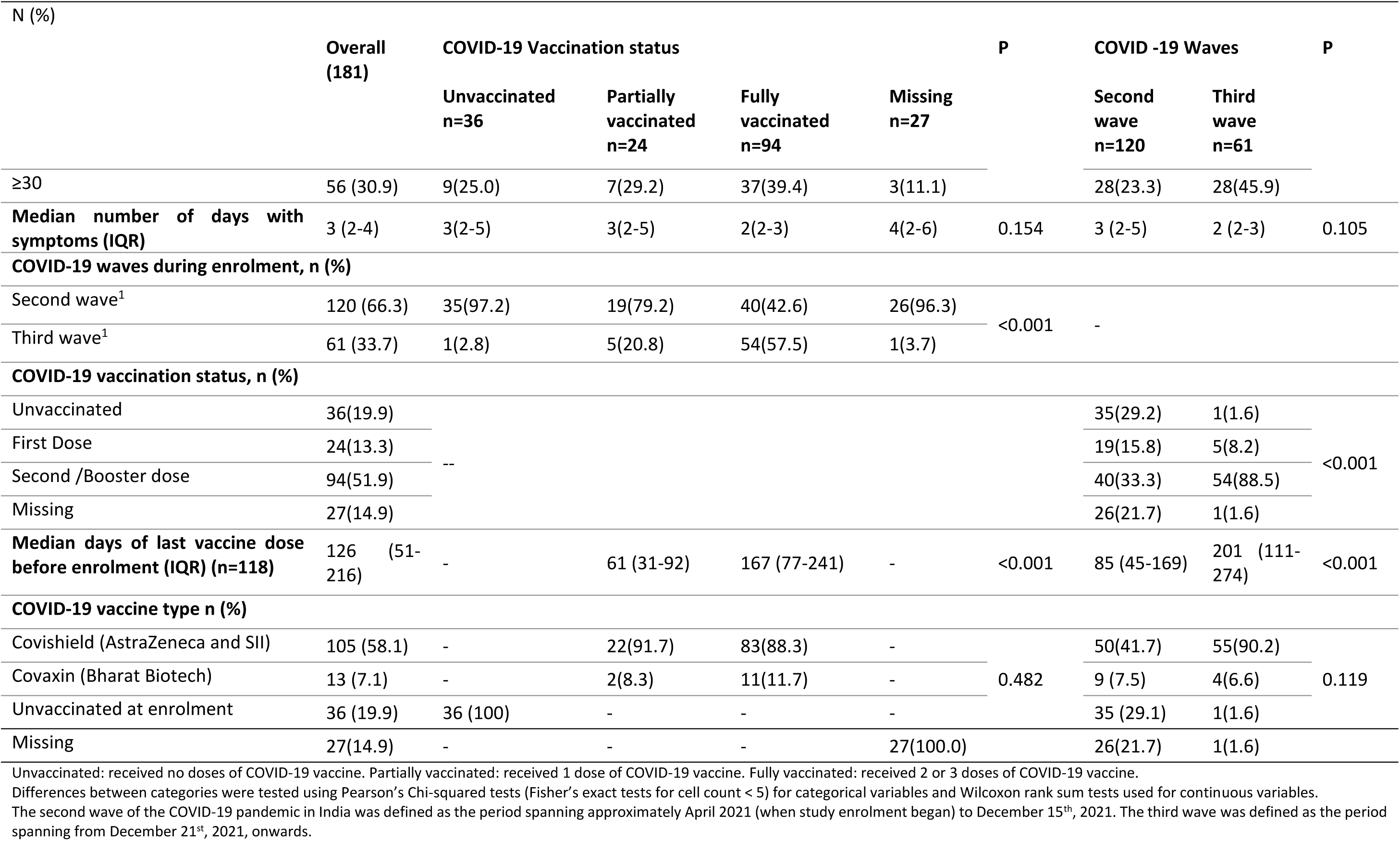
Baseline characteristics of enrolled participants (N=181).

### Baseline characteristics of the enrolled participants across exposure variables (vaccination status and waves of COVID-19 pandemic in India)

At the time of enrolment, one-fifth (19.9%) of individuals were unvaccinated, 13.3% had their first dose of COVID-19 vaccine, and 51.9% patients enrolled were fully vaccinated with two or three doses; 14.9% did not provide information on their vaccination status. The median days since last vaccine dose before enrolment was 126 days (IQR: 51-216). Fifty-eight percent individuals received COVISHIELD vaccine by AstraZeneca and Serum Institute of India, 7.1% received COVAXIN (Bharat Biotech) which was approved and introduced later, 19.9% were unvaccinated at the time of enrolment, and rest around 15% did not provide their vaccination details (Table 1).

A greater proportion of unvaccinated and fully vaccinated individuals was male (*P=*0.049). Additionally, a greater proportion of partially vaccinated individuals was in 30-44 years age group (*P=*0.008), had graduate education (*P=*0.006), was enrolled during the second wave (*P<*0.001), and was using any COVID-19 medications (*P=*0.061). Full vaccination was common in older participants who were more than 44 years, and among those who reported hypertension (*P=*0.023), any pre-existing conditions, and obesity. The median days from last dose before enrolment in partially vaccinated participants was 61 days (IQR: 31- 92) while it was 167 ([range 77-241] days for the fully vaccinated group (*P<*0.001]) (Table 1).

Most (66.3%) of the study participants were enrolled during the second wave. A larger proportion of participants who enrolled in the second wave were aged 30-44 years (*P=*0.381), and had normal BMI compared to third wave participants (*P=*0.019). All participants enrolled in the third wave had secondary or higher education whereas about one tenth of those enrolled in the second wave had primary or less education (*P=*0.004). Close to half (46%) of participants enrolled in the third wave were obese, compared with 23.3% of participants enrolled in the second wave (Table 1).

### Levels of inflammatory and immunological biomarkers of COVID-19 infected participants across vaccination status and waves of COVID-19 (unadjusted analyses)

We first compared unadjusted median biomarker levels across vaccination status at enrolment and the COVID-19 wave in which the participants were enrolled. The unadjusted median CRP, LDH, ferritin, D-dimer and IL-6 levels were lower in fully and partially vaccinated groups as compared to the unvaccinated group (*P<*0.001 for LDH and IL-6, and *P=* 0.005 for ferritin). The unadjusted levels of other inflammatory markers Ang-2 and sTREM-1 were higher in both vaccinated groups than unvaccinated group, but not significant (*P=*0.454 and *P=*0.977 respectively). Unadjusted IgG levels were significantly elevated in vaccinated groups as compared to the unvaccinated group especially in partially vaccinated people (*P=*0.009), while unadjusted IgM levels were also notably high in partially vaccinated group and lower in fully vaccinated people (*P=*0.014) (Table 2).

**Table 2.**
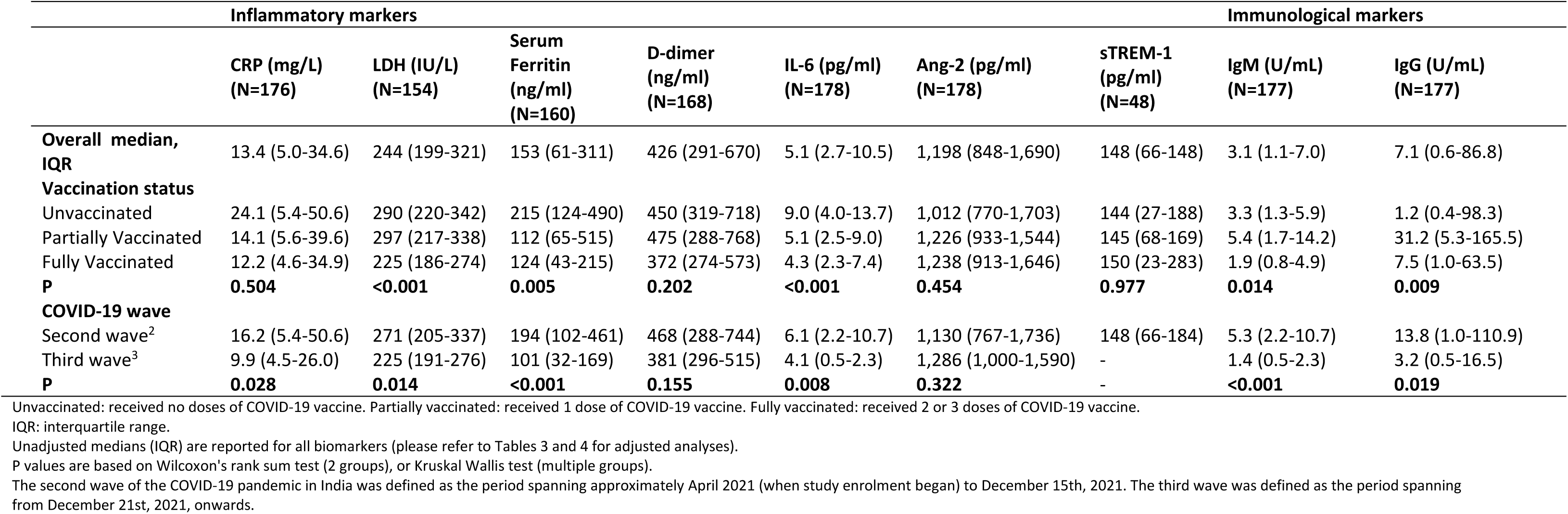
Median (interquartile range) levels of inflammatory and immunological biomarkers overall and across participant characteristics.

|  | Inflammatory markers |  |  |  |  |  | Immunological markers |  |  |
| --- | --- | --- | --- | --- | --- | --- | --- | --- | --- |
|  | CRP (mg/L)<br>(N=176) | LDH (IU/L)<br>(N=154) | Serum<br>Ferritin<br>(ng/ml)<br>(N=160) | D-dimer<br>(ng/ml)<br>(N=168) | IL-6 (pg/ml)<br>(N=178) | Ang-2 (pg/ml)<br>(N=178) | sTREM-1<br>(pg/ml)<br>(N=48) | IgM (U/mL)<br>(N=177) | IgG (U/mL)<br>(N=177) |
| **Overall median, IQR** | 13.4 (5.0-34.6) | 244 (199-321) | 153 (61-311) | 426 (291-670) | 5.1 (2.7-10.5) | 1,198 (848-1,690) | 148 (66-148) | 3.1 (1.1-7.0) | 7.1 (0.6-86.8) |
| **Vaccination status** |  |  |  |  |  |  |  |  |  |
| Unvaccinated | 24.1 (5.4-50.6) | 290 (220-342) | 215 (124-490) | 450 (319-718) | 9.0 (4.0-13.7) | 1,012 (770-1,703) | 144 (27-188) | 3.3 (1.3-5.9) | 1.2 (0.4-98.3) |
| Partially Vaccinated | 14.1 (5.6-39.6) | 297 (217-338) | 112 (65-515) | 475 (288-768) | 5.1 (2.5-9.0) | 1,226 (933-1,544) | 145 (68-169) | 5.4 (1.7-14.2) | 31.2 (5.3-165.5) |
| Fully Vaccinated | 12.2 (4.6-34.9) | 225 (186-274) | 124 (43-215) | 372 (274-573) | 4.3 (2.3-7.4) | 1,238 (913-1,646) | 150 (23-283) | 1.9 (0.8-4.9) | 7.5 (1.0-63.5) |
| **P** | **0.504** | **<0.001** | **0.005** | **0.202** | **<0.001** | **0.454** | **0.977** | **0.014** | **0.009** |
| **COVID-19 wave** |  |  |  |  |  |  |  |  |  |
| Second wave<sup>2</sup> | 16.2 (5.4-50.6) | 271 (205-337) | 194 (102-461) | 468 (288-744) | 6.1 (2.2-10.7) | 1,130 (767-1,736) | 148 (66-184) | 5.3 (2.2-10.7) | 13.8 (1.0-110.9) |
| Third wave<sup>3</sup> | 9.9 (4.5-26.0) | 225 (191-276) | 101 (32-169) | 381 (296-515) | 4.1 (0.5-2.3) | 1,286 (1,000-1,590) | - | 1.4 (0.5-2.3) | 3.2 (0.5-16.5) |
| **P** | **0.028** | **0.014** | **<0.001** | **0.155** | **0.008** | **0.322** | - | **<0.001** | **0.019** |

Unvaccinated: received no doses of COVID-19 vaccine. Partially vaccinated: received 1 dose of COVID-19 vaccine. Fully vaccinated: received 2 or 3 doses of COVID-19 vaccine.
IQR: interquartile range.
Unadjusted medians (IQR) are reported for all biomarkers (please refer to Tables 3 and 4 for adjusted analyses).
P values are based on Wilcoxon's rank sum test (2 groups), or Kruskal Wallis test (multiple groups).
The second wave of the COVID-19 pandemic in India was defined as the period spanning approximately April 2021 (when study enrolment began) to December 15th, 2021. The third wave was defined as the period spanning from December 21st, 2021, onwards.

The study participants enrolled in the second wave had a higher median level of unadjusted CRP, LDH, ferritin, and IL-6 compared to individuals enrolled in the third wave (*P<*0.05 for all). These participants also had lower unadjusted median D-dimer levels, though this was not significant (*P=*0.155). Furthermore, unadjusted plasma levels of IgM and IgG antibodies were significantly lower in the third wave compared with second wave participants (*P<*0.001 and *P=*0.019 respectively) (Table 2).

### Regression analysis of effect of vaccination status and wave of COVID-19 pandemic on inflammatory and immunological biomarkers (adjusted analyses)

In fully-adjusted multivariable regression models, full vaccination (but not partial vaccination) was associated with lower LDH and Ang-2; LDH (β: −77.4, 95% CI: −89.3 – −65.4, *P<*0.001) and Ang-2 (β: −61.0, 95%CI: −121.0 – −1.0, *P=*0.046) respectively. Full vaccination was also associated with lower D-dimer levels, but the association did not reach statistical significance (β: −248.9, 95% CI:-1,009.4 – 511.7, *P=*0.521). Furthermore, partial and full vaccination was associated with lower IL-6 levels as compared to no vaccination, with similar magnitudes of difference for both groups but significant associations only for the partially vaccinated group (β: −41.8, 95% CI: −81.7 – −1.9, *P=*0.040). In a subset of the cohort, in fully vaccinated participants had higher sTREM-1 levels (β:72.6, 95% CI: 70.6–74.5, *P<*0.001) (Table 3).

**Table 3.** Estimates of association between COVID-19 vaccination status and inflammatory and immunological biomarkers.

|  | **β Coefficient (95% Conf. Interval)** | **P** |
| --- | --- | --- |
| **CRP (mg/L)** |  |  |
| Partially vaccinated | -26.1 (-102.1-50.0) | 0.502 |
| Fully Vaccinated | -28.5 (-88.0-30.9) | 0.347 |
| **LDH (IU/L)** |  |  |
| Partially vaccinated | 9.3 (-31.7-50.2) | 0.658 |
| Fully Vaccinated | -77.4 (-89.3--65.4) | <0.001 |
| **Serum Ferritin (ng/ml)** |  |  |
| Partially vaccinated | -38.7 (-439.6-362.2) | 0.850 |
| Fully Vaccinated | -177.4 (-691.3-336.5) | 0.499 |
| **D-Dimer (ng/ml)** |  |  |
| Partially vaccinated | 95.4 (-450.3-641.2) | 0.732 |
| Fully Vaccinated | -248.9 (-1,009.4-511.7) | 0.521 |
| **IL-6 (pg/ml)** |  |  |
| Partially vaccinated | -41.8 (-81.7--1.9) | 0.040 |
| Fully Vaccinated | -36.3 (-81.4-8.9) | 0.116 |
| **Angiopoietin-2 (pg/ml)** |  |  |
| Partially vaccinated | 1.5 (-379.9-382.9) | 0.994 |
| Fully Vaccinated | -61.0 (-121.0--1.0) | 0.046 |
| **sTREM-1 (pg/ml)** |  |  |
| Partially vaccinated | 16.5 (-91.4-124.3) | 0.765 |
| Fully Vaccinated | 72.6 (70.6-74.5) | <0.001 |
| **IgM (U/mL)** |  |  |
| Partially vaccinated | 52.7 (-145.7-251.0) | 0.603 |
| Fully Vaccinated | -13.9 (-43.6-15.9) | 0.360 |
| **IgG (U/mL)** |  |  |
| Partially vaccinated | 61.5 (48.9-74.1) | <0.001 |
| Fully Vaccinated | 62.2 (56.7-67.6) | <0.001 |

Unvaccinated: received no doses of COVID-19 vaccine. Partially vaccinated: received 1 dose of COVID-19 vaccine. Fully vaccinated: received 2 or 3 doses of COVID-19 vaccine.
95% CI: 95% confidence interval.
Estimates based on mixed effect regression models with robust standard errors.
The comparison group in the model is unvaccinated participants.
The model is adjusted for wave enrolled, age, gender, type of vaccine, BMI, COVID medications, pre-existing diseases, days of symptoms, health behaviour (drinking and smoking), and clustered by hospital sites.

Although not significant, decreasing trends in CRP in partially vaccinated (β: −26.1, 95% CI: - 102.1 – 50.0, *P=*0.502) and fully vaccinated individuals (β: −28.5, 95% CI: −88.0 – 30.9, *P=*0.347) were observed, similarly decreasing ferritin levels in partial and fully vaccinated participants were also noticed (β: −38.7, 95% CI: −439.6– 362.2, *P=*0.850 and β: −177.4, 95% CI: −691.3 – 336.5, *P=*0.499 respectively) (Table 3).

Significant associations between partial and full vaccination and elevated IgG were detected (β: 61.5, 95% CI: 48.9 – 74.1, *P<*0.001, and β: 62.2, 95% CI: 56.7 – 67.6, *P<*0.001 respectively). Estimates suggested increased levels of IgM associated with partial vaccination. However, these estimates of increase were not significant, and were also not statistically different from estimates for fully-vaccinated individuals, with fully overlapping confidence intervals (Table 3).

Additionally, in these fully-adjusted analyses, enrolment in the third wave was associated with significantly lower IL-6 (β: −12.0, 95% CI: −12.6 – −11.4, *P<*0.001), and CRP which was borderline significant (β: −21.7, 95% CI: −44.0 – 0.6, *P=* 0.056). On the other hand, higher D- dimer values were significantly associated with enrolment in third-wave (β: 322.5, 95% CI: 80.9 – 564.2, *P=*0.009). Other inflammatory markers including ferritin, LDH, and Ang-2 levels were also lower in third-wave versus second-wave participants, but differences in these biomarkers were not significant (Table 4).

**Table 4.** Estimates of association between COVID −19 waves and inflammatory and immunological biomarkers.

| | $\beta$ Coefficient (95% Conf. Interval) | P |
| --- | --- | --- |
| **CRP (mg/L)** |  |  |
| Enrolled wave 3 | -21.7 (-44.0-0.6) | 0.056 |
| **LDH (IU/L)** |  |  |
| Enrolled wave 3 | -14.4 (-30.8-1.9) | 0.084 |
| **Serum ferritin (ng/ml)** |  |  |
| Enrolled wave 3 | -29.1 (-171.1-112.9) | 0.688 |
| **D-Dimer (ng/ml)** |  |  |
| Enrolled wave 3 | 322.5 (80.9-564.2) | 0.009 |
| **IL-6 (pg/ml)** |  |  |
| Enrolled wave 3 | -12.0 (-12.6--11.4) | <0.001 |
| **Angiopoietin-2 (pg/ml)** |  |  |
| Enrolled wave 3 | -26.5 (-139.1-86.0) | 0.644 |
| **sTREM-1 (pg/ml)** |  |  |
| Enrolled wave 3 | -- | -- |
| **IgM (U/mL)** |  |  |
| Enrolled wave 3 | -23.6 (-42.3--4.9) | 0.013 |
| **IgG (U/mL)** |  |  |
| Enrolled wave 3 | -77.6 (-99.4--55.8) | <0.001 |

95% CI: 95% confidence interval.
Estimates based on mixed effects regression models with robust standard errors
The comparison group is wave 2 participants
The model is adjusted for age, gender, vaccination status (unvaccinated: 0 doses of COVID-19 vaccine, partially vaccinated: 1 dose, fully vaccinated: 2 or 3 doses) and type of vaccine, BMI, COVID medications, pre-existing diseases, days of symptoms, health behaviour (drinking and smoking), and clustered by hospital sites.
The second wave of the COVID-19 pandemic in India was defined as the period spanning approximately April 2021 (when study enrolment began) to December 15th, 2021. The third wave was defined as the period spanning from December 21st, 2021, onwards.

Significant lower IgG and IgM were associated with enrolment in the third wave as compared to second-wave; IgM (β: −23.6, 95% CI: −42.3 – −4.9, *P=* 0.013) and IgG (β: −77.6, 95% CI: −99.4 – −55.8, *P<*0.001) (Table 4).

## Discussion

In this study, we found that levels of inflammatory markers, particularly LDH and Ang-2 were significantly lower in fully vaccinated participants compared to unvaccinated participants with COVID-19. Furthermore, we observed some evidence of lower IL-6 levels in both partially and fully vaccinated participants, and substantially elevated IgG levels in both partially and fully vaccinated participants. We also found that infection during the third wave was associated with lower CRP, IL-6, and IgG levels, and higher D-Dimer levels.

Inflammation is a defence mechanism that is activated by a coordinated signalling pathway, and also regulates inflammatory mediator levels in host tissue cells and inflammatory cells recruited from the blood.^[15]^ Previous studies have shown that vaccinated patients have reduced severity of COVID-19 disease compared to unvaccinated patients, and individuals with different vaccination doses have variable immune-inflammatory characteristics including the existence of higher neutralizing antibodies response and lower inflammatory markers.^[16,17]^

CRP LDH, ferritin, D-dimer, and IL-6 are inflammatory markers explored commonly in COVID- 19 and other research studies. However, evidence on the association between COVID-19 vaccination and variants and other inflammatory markers such as Ang-2 and sTREM-1, as performed in our study, is scarce – we were unable to find any relevant studies during our search of the literature on these associations. Some previous studies, including from South Asia, have examined the relation between COVID-19 vaccination or variants and commonly measured inflammatory and immunological markers.^[10,18,19]^ Similar to our study, data from these previous reports suggest generally inverse associations between vaccination against COVID-19 and commonly measured inflammatory markers, and positive associations between vaccination and immunological markers^[10,18,19]^. For example, an observational study conducted in Pakistan on a cohort of 434 COVID-19 patients reported significantly higher LDH, ferritin, and D-dimer levels in unvaccinated individuals compared to vaccinated individuals.^[18]^ In another study comparing unvaccinated COVID-19 patients with the fully vaccinated group, significantly lower levels of IL-6 along with CRP and D-dimer were reported in the fully vaccinated group. This study also showed noticeably higher IgG in fully vaccinated individuals.^[19]^ Similarly, significantly higher IgG levels among partially vaccinated individuals were also observed in our study.

Also corroborating our results, a recent observational study conducted in India on 1160 hospitalized COVID-19 patients found similar trends of immunological and inflammatory biomarkers. This included raised D-Dimer levels reported after the first vaccination, but lower levels after 2 or 3 doses, and non-significant declining CRP.^[10]^ Taken together with these previous data, the lower CRP, LDH, ferritin, D-dimer, Ang-2, and IL-6 inflammatory mediators among vaccinated individuals in our study add to the evidence that vaccine- induced protection with higher immunological mediators may help to reduce inflammation, which has been associated with more severe clinical outcomes in COVID-19.^[20]^

Previous evidence also suggests that the SARS-CoV-2 Omicron variant is less severe compared to other variants, and limited data from previous reports suggest that the Omicron variant may induce a smaller inflammatory and immune response.^[8,21,22]^ The possible reasons noted are less effective replication in the lung parenchyma,^[23]^ and sustained conservation of T-cell mediated immunity from previous infection or vaccination.^[24]^ A study conducted in Australia assessed severity of COVID-19 variants through laboratory findings, and observed lower inflammatory markers, particularly ferritin, LDH, and CRP levels, linked with Omicron variant compared to Delta variant. This study also found no significant differences in D-dimer levels.^[8]^ A recent comparative study of the three waves of COVID-19 in India reported that infection with the Omicron variant versus Delta variant was associated with significantly lower levels of CRP and IL-6.^[21]^ Another study conducted in Bangladesh reported higher levels of IgG, and higher risk of hospitalization and oxygen support were associated with infection with the Delta compared to Omicron variant.^[22]^ Contributing to growing evidence suggesting that the Omicron variant is associated with less severe disease, our study also suggests that participants enrolled in waves of infection corresponding to the Omicron variant had lower inflammatory and immunological biomarkers.

Contrary to other inflammatory biomarkers, D-dimer concentration in our study was substantially higher in the third wave compared to the second wave participants. A study on D-dimer levels suggested that early management of infection with steroids and anticoagulants in the second wave may affect the production of D-dimer and limit the endothelial damage, however, only a few participants were on steroids and anticoagulants in our study.^[25]^ Other reasons could be greater proportion of elderly individuals and high BMI reported in the third wave in our study which are independent risk factors of high D-dimer levels.^[26,27]^ Importantly, we adjusted for these factors in analyses, suggesting that other unmeasured clinical factors may underlie the observed associations.

The immune response is a dynamic process that might also change the inflammatory parameters. Inflammatory response increases rapidly during the initial phase of infection and falls during the recovery period, therefore, longitudinal changes in inflammatory markers are more important rather than values of biomarkers at a single time point.^[28]^

Typically, after the onset of symptoms, immunological markers IgM and IgG can be detected during the first week and at around two weeks respectively,^[29]^ and gradually these antibodies decline with time. The current study suggests that antibodies were higher among vaccinated participants who later became ill with COVID-19 compared with unvaccinated individuals. Particularly IgG antibodies remained high even after receiving two or three vaccine doses compared to those who did not receive any vaccine doses. Together with recent evidence from India indicating the effect of IgG levels on COVID-19-related measures such as viral emission^[30]^ these findings help characterize the biological effects of COVID-19 vaccination that may underlie disease severity and outcomes among SARS-CoV-2-infected individuals.

This study has a few limitations. Information on prior COVID-19 infection was not collected from the study participants which could affect in the vaccine-induced immune response and its impact on inflammatory biomarkers. Secondly, different types of COVID-19 vaccines can elicit diverse antibody responses, although in this study most (89%) of participants had received the adenoviral vector vaccine (COVISHIELD) in comparison to the whole cell inactivated virus (COVAXIN). Additionally, genome sequencing for confirmation of COVID-19 variants was not performed in this study, and classification of two waves of the COVID-19 pandemic was made based on defined time points by the federal COVID-19 task forces and previous research articles.^[14]^ Therefore, there is the possibility that participants enrolled at beginning of the third wave may have been infected with either the Delta or the Omicron variant of the SARS-CoV-2 virus. Analyses were based on a small sample size, particularly for unvaccinated individuals in the third wave – which was likely due to good uptake of vaccines during the study period. This may have limited our ability to detect associations in certain cases, as evidenced by the wide confidence intervals for linear regression estimates.

Additionally, this study was based among individuals with mild-to-moderate COVID-19 symptoms and no severe outcomes, meaning that we were unable to clearly examine the effect of vaccination status, wave of enrolment, and biomarker levels on clinical outcomes.

The foremost strength of this study is the diverse set of inflammatory and immunological biomarkers of COVID-19 infection explored, which gives an opportunity to understand the biological changes related to infection that are affected by prior COVID-19 vaccination and by SARS-CoV-2 variants. Furthermore, we were able to account for a range of potential demographic, clinical and health-related confounders by adjusting for these in final regression models. Changes in trends cross unadjusted and adjusted analyses suggested the importance of accounting for these characteristics when drawing conclusions regarding the potential effect of vaccination status and COVID-19 wave on biomarker levels. Additionally, this study also provides indicative data on the different clinical manifestations of two SARS-CoV-2 variants of concern. The study setting in India enables applicability of findings to the wider South Asia context where evidence on the effect of COVID-19 vaccine and SARS-CoV-2 variants on inflammatory and immunological responses to SARS-CoV-2 infection remains scarce.

## Conclusion

This study indicates that the inflammatory and immunologic response to SARS-CoV-2 infection may be least partly associated with prior COVID-19 vaccination status and with the SARS-CoV-2 variant. This study provides data outlining potential pathways through which COVID-19 vaccination may protect against, and SARS-CoV-2 variants may influence, the likelihood of severe disease.

## Competing interests

All authors declare no conflicts of interest.

## Registration

Clinical Trial Registry of India (CTRI): CTRI/2021/04/032593 (https://ctri.nic.in/Clinicaltrials/pmaindet2.php?EncHid=NTQ1MTA=&Enc=&userName=CTRI/2021/04/032593)

## Ethical approval

This study was conducted in accordance with the Declaration of Helsinki, and informed written consent was taken from study participants. Ethical approval was obtained from the Institutional Review Board of the Harvard T.H. Chan School of Public Health (Protocol No. IRB20-1425), the University Health Network Research Ethics Board (20-5775), the Institutional Research Ethics Committee of the Foundation for Medical Research (IREC No. FMR/IREC/C19/02/2020), the Institutional Review Board of Saifee Hospital (Project No. EC/008/2020) and the KEM Hospital Research Centre Ethics Committee (KEMHRC ID No. 2027).

## Author contributions

KKS, UP, WWF, KCK, YD, and NM conceptualized the study along with YM, SS, PD, GG and MW. KKS, YD, YM, SS, PD and GG were involved in data acquisition, and in study monitoring along with KCK, WWF, NM and UP. MW provided statistical expertise. KKS and UP analysed the data, KKS drafted the first draft of manuscript, and all authors reviewed and critically revised the draft and approved the final manuscript.

## Data Availability

All data produced in the present study are available upon reasonable request to the authors

## Notes

### Competing Interest Statement

The authors have declared no competing interest.

### Clinical Trial

NCT04641195; CTRI/2021/04/032593

### Clinical Protocols

https://bmjopen.bmj.com/content/12/8/e061301.abstract

### Funding Statement

The collection of baseline data was done under a randomized control trial that was supported by the Canadian Institutes of Health Research, Operating Grant: COVID-19 Rapid Research Funding OpportunityTherapeutics (application number: 447092), and the Canada Research Chair program (to KCK). The funding bodies had no role in study design and procedures, or the decision to submit manuscripts for publication.

### Summary of Updates

Only author's spelling has been edited rest are the same.

